# Attitudes and Perceptions of Medical Researchers Towards the Use of Artificial Intelligence Chatbots in the Scientific Process: A Protocol for a Cross-Sectional Survey

**DOI:** 10.1101/2023.07.26.23293211

**Authors:** Jeremy Y. Ng, Sharleen G. Maduranayagam, Cynthia Lokker, Alfonso Iorio, R. Brian Haynes, David Moher

**Author notes:** Corresponding Author Address: The Ottawa Hospital, General Campus, Centre for Practice Changing Research Building, 501 Smyth Road, PO BOX 201B, Ottawa, ON, K1H 8L6 Canada.

## Abstract

Artificial intelligence (AI) refers to computer systems or robots that can perform tasks associated with human intelligence, such as reasoning, problem-solving, and learning. While AI programs have not matched human versatility, they are increasingly used in various domains like self-driving cars, speech transcription, medical diagnosis, and smart assistants. AI has benefited fields like medicine, healthcare, and scientific research by improving productivity, reducing errors, and lowering costs. AI chatbots are conversational programs used for customer service, mental health support, and education. In scientific research, chatbots have the potential to automate tasks like literature searches, data analysis, and manuscript writing, improving efficiency and addressing the reproducibility crisis. However, there are challenges to overcome, including accuracy, reliability, ethical concerns, and limitations of current chatbot models. Scholarly publishing faces debates about authorship and guidelines have been established by journals and publishing organizations regarding the use of AI chatbots. To understand researchers’ attitudes towards AI chatbots, an international survey is proposed to explore their familiarity, perceived benefits, limitations, and factors influencing adoption. Findings can guide policy development and implementation of AI chatbots in scientific research.

## Background

Artificial intelligence (AI) broadly refers to the ability of a computer or a computer-controlled robot to perform tasks typically associated with human intelligence and its cognitive functions (Copeland, 2023; Petersson et al., 2022; Schroer, n.d.). These include the ability to reason, discover meaning, problem solve, generalize, and/or learn from past experiences (Copeland, 2023). Although AI programs have not yet achieved the versatility of human intelligence, specialized AI programs are increasingly becoming a part of daily life across many domains (Schroer, n.d.). Examples include self-driving cars, transcribing speech, medical diagnoses, search engines, and perhaps more popularly, smart assistants such as Siri and Alexa (Copeland, 2023; Nadarzynski et al., 2019; Schroer, n.d.). Generally, AI usage in these fields has been beneficial by improving productivity and efficiency (Schroer, n.d.). For example, in medicine and healthcare, AI systems have helped reduce diagnostic and therapeutic errors, promoted and increased physical activity, and are estimated to have reduced healthcare costs (Ali et al., 2023; Nadarzynski et al., 2019; Palanica et al., 2019). It is unsurprising then that AI has also become an increasingly important tool in scientific research, particularly in fields where large amounts of data need to be analysed and interpreted.

Chatbots are AI programs designed to simulate conversation with human users through text or speech (Adamopoulou & Moussiades, 2020; Palanica et al., 2019). In response to prompts and questions, these chatbots can generate relevant responses. Some popular AI chatbots include ChatGPT, Bing Chat, YouChat, and Google Bard (Google, n.d.; Microsoft, n.d.; OpenAI, n.d.; You.com, n.d.). AI chatbots have been used for a variety of purposes, including customer service, mental health support, and education (Nadarzynski et al., 2019; Palanica et al., 2019). In scientific research, these chatbots have the potential to automate tasks such as literature searches and reviews, data analyses and interpretations of large datasets, experimental design, and manuscript writing (Nature, 2023; Sallam, 2023; Salvagno et al, 2023). They may also improve the readability of scientific articles through language reviews and could help bypass language related publishing barriers for non-native speakers, potentially helping increase equity and diversity in research (Liebrenz et al., 2023; Sallam, 2023; Salvagno et al., 2023). Overall, chatbots could improve the efficiency and accuracy of scientific research, as well as help to address the reproducibility crisis in science. AI systems can be trained to potentially differentiate between reproducible and non-reproducible studies by estimating a study’s likelihood of replication (Yang et al., 2020). As conducting manual replication research can be time-consuming and costly, AI systems may also help reduce these resource burdens on researchers (Yang et al., 2020), However, the use of AI chatbots in scientific research is still in its early stages, and there are several potential challenges and limitations that need to be addressed. Some major concerns include the accuracy and reliability of chatbots in performing or reporting scientific tasks, as well as ethical issues regarding the use of AI in research. For example, there are well-noted shortcomings of using ChatGPT, a popular AI chatbot accessible to the public, in scientific writing. These include its ability to generate seemingly plausible but ultimately inaccurate/incorrect content, citation errors, insufficient references, and referencing to non-existent sources, and its limited knowledge if the chatbot is trained on noncurrent datasets (Sallam, 2023). There are also ethical and legal issues associated with chatbot use in scientific research, such as plagiarism, a risk of amplifying and perpetuating biases and inaccuracies, the risk of research fraud, copyright issues, and the lack of transparency regarding content generation (Liebrenz et al., 2023; Sallam, 2023; Salvagno et al., 2023).

Considering these challenges and limitations, there is a potential for chatbots to also spread misinformation, with harmful consequences (Liebrenz et al., 2023; Sallam, 2023).

Regarding scholarly publishing, there is debate on whether AI chatbots should be recognized as authors (Lee, 2023; Liebrenz et al., 2023; Marchandot et al., 2023; Polonsky & Rotman, 2023; Stokel-Walker, 2023). In response to a rise in researchers listing ChatGPT as a co-author on their manuscripts, many journals’, including *Nature* and the *Lancet*, have stated that Large Language Models (LLMs) (e.g., ChatGPT) are not permitted to be listed as a co-author and authors must disclose any use of text-generating tools in submitted manuscripts (Brainard, 2023; Lee, 2023). Publisher organizations/associations, such as the Committee on Publication Ethics (COPE) and the World Association of Medical Editors (WAME) have echoed the same guidelines, stating that AI tools (i.e., chatbots) do not satisfy their authorship criteria as they are unable to take responsibility for their scripts (Brainard, 2023; COPE, 2023; Zielinski et al., 2023). In addition to agreeing that AI tools cannot be authors, one journal, *Science,* has already stated that it will not accept any AI generated material (e.g., text, images, figures, etc.) in submitted manuscripts (AAAS, n.d.).

To better understand medical researchers’ attitudes and perceptions towards the use of AI chatbots in the scientific process, we propose an international cross-sectional survey. This survey will seek to investigate the extent to which researchers are familiar with AI chatbots, the perceived potential benefits and limitations of using these chatbots in scientific research, and the factors that may influence their adoption. This survey will be the first of this magnitude to provide an international perspective on researcher’s knowledge, use, and perceptions of AI chatbots in the scientific process.

By gaining a better understanding of researchers’ attitudes towards AI chatbots, our findings can help to inform the development and implementation of policies and uses of these tools in scientific research. Ultimately, the use of AI chatbots has the potential to revolutionize the way that scientific research is conducted, and this survey represents an important step towards understanding this potential.

## Methods

### Open Science Statement

Prior to beginning this study, we will apply for permission from the Ottawa Health Sciences Research Ethics Board (REB). This study protocol will be registered on the Open Science Framework (OSF) (https://osf.io) before participant recruiting begins. The study materials and data will be made available via OSF at the time of publication submission. The resulting manuscript will be posted as an OSF Preprint.

### Study Design

We will conduct an anonymous, cross-sectional closed survey of published medical researchers to investigate their attitudes and perceptions towards the use of AI chatbots in the scientific process. The survey will be administered online, using SurveyMonkey, a secure web-based survey tool. The survey will consist of both closed-ended (e.g., multiple choice, yes/no) and open-ended questions (i.e., participants will type their answer in the text box provided), covering the following topics:

- Demographic information: age, gender, country of employment, level of education, publication record, and years of experience in medical research.
- Experience with AI chatbots: extent to which participants are familiar with AI chatbots, their personal experience with AI chatbots, and their likelihood of utilizing chatbots in the future.
- Role of AI chatbots in the scientific process: participants’ perceptions of AI chatbot utilization in different steps of the scientific process, and the potential impact of chatbots on the future of scientific research.
- Perceived benefits of AI chatbots in the scientific process: participants’ perceptions of the potential benefits of using AI chatbots in scientific research, such as increased efficiency, reduced human workload, and increased accuracy and dissemination.
- Perceived challenges of AI chatbots in the scientific process: participants’ perceptions of the potential challenges of using AI chatbots in scientific research, such as ethical concerns, risk of bias, and potential inaccuracies.
- Open-ended questions: participants will have the opportunity to provide additional comments and feedback on the use of AI chatbots in scientific research.

### Sampling Framework

A complete list of all journals indexed in MEDLINE (approximately 5300 journals as of April 2023) will be obtained along with their NLM IDs. A search strategy of these NLM IDs will be developed in OVID MEDLINE and limited to the records indexed over the last two months at the time of searching. This period was chosen as researchers who have published between these dates are likely to still be actively involved and available to respond to a survey request. Authors of all article types will be included. Any duplicates will be removed prior to recruitment. PMID numbers associated with these articles from all records yielded will be exported from OVID as a .csv file and inputted into an R script (created based on the easyPubMed package) to capture author names, affiliated institutions, and email addresses. We will also use the “Find Full Text” function in EndNote to retrieve PDF files of these articles. These files will be run in another R script for text recognition to extract email addresses. All generated results will be combined into a master list and cleaned for potential errors/duplicates before survey administration. A search strategy template is provided in **Appendix 1**.

### Inclusion Criteria

Participants must meet the following inclusion criteria: 1) are self-identified medical researchers (of any kind, whereby their research in one way or another contributes to the field of medicine) who are a corresponding author on an article indexed in MEDLINE, 2) have completed at least one terminal degree in their respective field of study (e.g., PhD or equivalent in your respective field, MD or equivalent in your respective profession) or have >5 years of experience in a research-focused role (e.g., research coordinator), and 3) be fluent in English. Students (both undergraduate and graduate) are ineligible to participate in this survey.

### Participant Recruitment

Prospective participants will be recruited from a range of academic disciplines within the field of medicine. Participants must identify as a medical researcher and have published at least one article in the last 12 months in a MEDLINE-indexed journal. We will use convenience sampling to recruit participants, targeting medical researchers identified by our sampling framework. An email containing text from a recruitment script approved by the REB, detailing the study and its purpose, and a link to the survey will be sent through email using the Mail Merge software. When participants click on the survey link, they will be directed to a webpage containing an informed consent form. Participants must agree to the form in order to proceed to the survey. This survey will be closed, meaning only invited participants will be able to participate.

If participants do not respond to the original invitation email, reminder emails will be sent after the first, second, third, and fourth weeks following the initial invitation. Following the fourth and final email, a 2-week waiting period will commence to accommodate remaining interested participants before the survey is closed permanently. Participants will have a total of 6 weeks to complete the survey. This will be stated clearly in both the initial and reminder emails.

The invitational survey will be administered online using SurveyMonkey. Completion of the survey will also be treated as implied consent. Monetary compensation will not be offered to participants and there is no requirement at any time to participate. Participation is entirely voluntary and anonymous, questions can be skipped if one does not wish to respond, and participation can be withdrawn at any time during the survey simply by closing the browser.

### Sample Size

As our initial list of names and emails will likely contain duplicate email addresses and names, and non-functioning emails, we anticipate emailing about 40 000 researchers after deduplication. This estimate was calculated using the following steps: approximately 5300 journals will be selected and PMID numbers associated with articles published in those journals over the last 2-month period at the time of searching will be retrieved. Using the assumption that at least 1 PMID number per journal per month is retrieved in this period, 120 000 PMID numbers will be retrieved from MEDLINE. After inputting the 120 000 PMIDs into R and operating on the assumption that every 3 PMIDs will yield at least 1 name/email address, an estimated 40 000 unique emails/names will be obtained. Assuming a 3-5% response rate, we can predict a minimum of 1200 survey responses being received. This sample size should provide sufficient statistical power to detect meaningful differences (if any) in attitudes and perceptions between different subgroups of participants (e.g., age, publication record, experience with AI chatbots). For example, since we are collecting information on participant ages, it may be possible to detect statistically significant differences in views on AI chatbot use in research between different age groups.

### Survey Instrument

The complete survey can be found in **Appendix 2**. The survey was created, distributed, and collected using the University of Ottawa’s version of the SurveyMonkey software (https://www.surveymonkey.com/). The survey instrument was developed based on a review of the literature and input from experts in the field of AI and scientific research. Survey questions were beta-tested by a team of authors prior to administration of the survey.

The survey will begin with a screening question, asking researchers if they identify as a medical researcher and have obtained at least one terminal degree. This will be followed by 7 demographic questions, asking participants about their current position, research area, gender, age, and country of employment. Then participants will be asked 8 questions about their familiarity and experience with AI chatbots, followed by 8 questions regarding the role of AI chatbots in the scientific process, and 4 questions about participants perceived benefits and challenges of using AI chatbots in research. This survey will conclude with an open-ended question for participants to offer feedback and additional thoughts regarding AI chatbot use in scientific research. The survey will contain 28 questions in total and will take approximately 15 minutes to complete.

### Data Management and Analysis

All responses will be collected using SurveyMonkey. The survey data will be exported and analysed using Microsoft Excel. Descriptive statistics, such as counts and percentages, will be generated and used to summarize the data collected in the survey. The demographic information of the participants will be compared and studied as well. As exploratory analysis, Chi-Square Crosstabs tests may be used if sufficient responses are obtained, to determine whether there are differences in attitudes and perceptions between different subgroups of participants (e.g., senior researchers versus early career researchers). Qualitative data collected through open-ended questions will be analysed thematically by two authors. Prior to beginning analysis, pilot coding will be conducted. Each author will individually code the responses of the first three survey respondents, and then collaborate to develop a shared code based on their results. After reaching a consensus on the code(s), they will thematically group all responses into the determined categories. These will be further defined and described for reporting into distinct tables.

### Ethical Considerations

We will obtain approval from the Ottawa Hospital Research Institute Research Ethics Board prior to conducting this study. Informed consent will be obtained from all participants prior to their participation in the survey. Participation in the survey will be voluntary, and participants will have the right to withdraw from the study at any point prior to submitting their completed survey. All data collected will be kept confidential and anonymous, and no identifying information will be collected. Therefore, once the survey is submitted, participants will not be able to withdraw from the study as their responses will be collected without any personal identifiers.

## Discussion

The main purpose of this study is to collect the attitudes and perceptions of medical researchers on the use of AI chatbots in the scientific process and their potential impact on how scientific research is conducted at a time when these chatbots are newly being applied to the field. AI chatbots can automate time-consuming tasks in the research process, provide writing and editing assistance, and potentially enhance research equity by helping non-native speakers bypass language barriers (Sallam, 2023). Although the use of AI chatbots in research is an exciting prospect, there are significant concerns and challenges regarding their usage that need urgent attention. For one, a code of ethics concerning the use of AI tools, such as chatbots, in academia and research is needed (Sallam, 2023; Salvagno et al., 2023). In order to begin developing and implementing AI use in research, it is necessary to understand how researchers perceive, think, and feel about using AI chatbots in the scientific process.

The results of this survey may serve to inform the development and implementation of AI tools in scientific research and policies surrounding their usage, considering the potential impact improper use may have on researchers, institutions, education, and the publishing industry, and the potential benefits appropriate use can offer towards the dissemination of knowledge.

### Strengths and Limitations

As we are utilizing a cross-sectional survey in this study, we are only observing our population of interest at one point in time and will not continue to follow them through time. This makes this survey type relatively inexpensive and quick and easy to conduct. As this survey aims to assess the thoughts of a large sample of researchers, its results are also generalizable to medical researchers. These researchers, stemming from different disciplines, will likely have varying opinions regarding AI chatbot use in scientific research. A final strength of our study is that we will only be retrieving names and email addresses of researchers from the most recent 2 months. This will minimize the possibility of bounce back/inactive emails. One limitation of our study design is that non-English speaking researchers will largely be excluded from our sample due to our own language and resource limitations. In addition, researchers who are only partially fluent in English may find some difficulty in participating in our survey. This would exclude their perceptions and attitudes; thus, our findings may not be applicable to those who primarily publish their research in languages other than English. Another limitation is that we will be underestimating the response rate as we cannot account for bounce back/inactive email accounts and autoreplies. A final limitation includes recall and non-response bias, which are inherent to the cross-sectional survey study design.

## List of Abbreviations

AI: artificial intelligence
ChatGPT: Chat Generative Pre-Trained Transformer
COPE: Committee on Publication Ethics
LLM: large language model
OSF: Open Science Framework
WAME: World Association of Medical Editors

## Declarations

### Ethics Approval and Consent to Participate

Ethics approval will be sought through the local research ethics board at the Ottawa Hospital Research Institute.

### Consent for Publication

All authors consent to this manuscript’s publication.

### Availability of Data and Materials

Raw data associated with this manuscript weill be made publicly available for download on Open Science Framework: https://doi.org/10.17605/OSF.IO/25Y8Q

### Authors’ Contributions

JYN: designed and conceptualized the study, drafted the manuscript, and gave final approval of the version to be published.

SM: made critical revisions to the manuscript, and gave final approval of the version to be published.

CL: made critical revisions to the manuscript, and gave final approval of the version to be published.

AI: made critical revisions to the manuscript, and gave final approval of the version to be published.

RBH: made critical revisions to the manuscript, and gave final approval of the version to be published.

DM: made critical revisions to the manuscript, and gave final approval of the version to be published.

## Data Availability

All data produced in the present work are contained in the manuscript, as this is a study protocol.

https://doi.org/10.17605/OSF.IO/25Y8Q

## Appendix 1: OVID MEDLINE Search Strategy Derived from All Journals Currently Indexed in MEDLINE

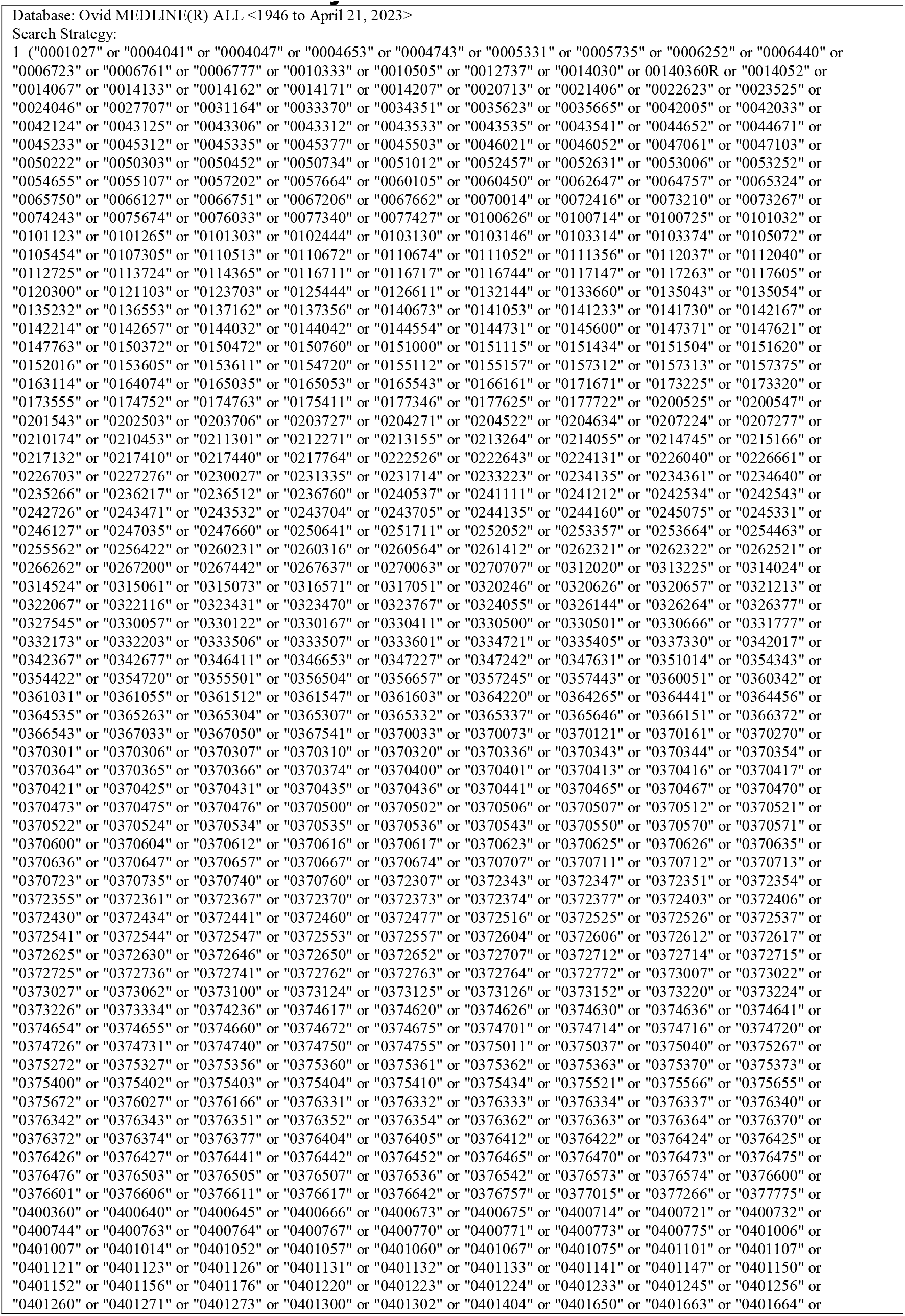
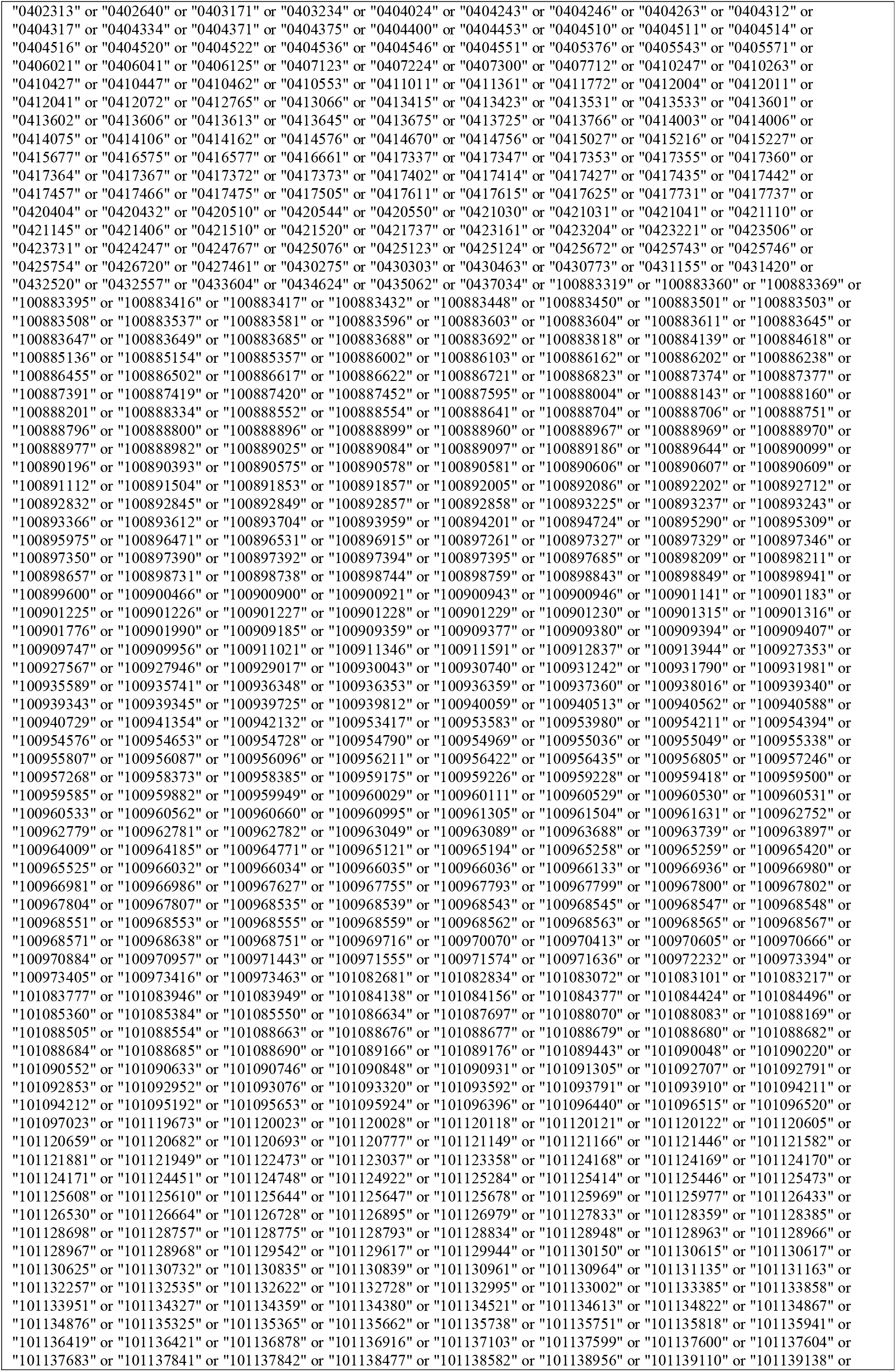
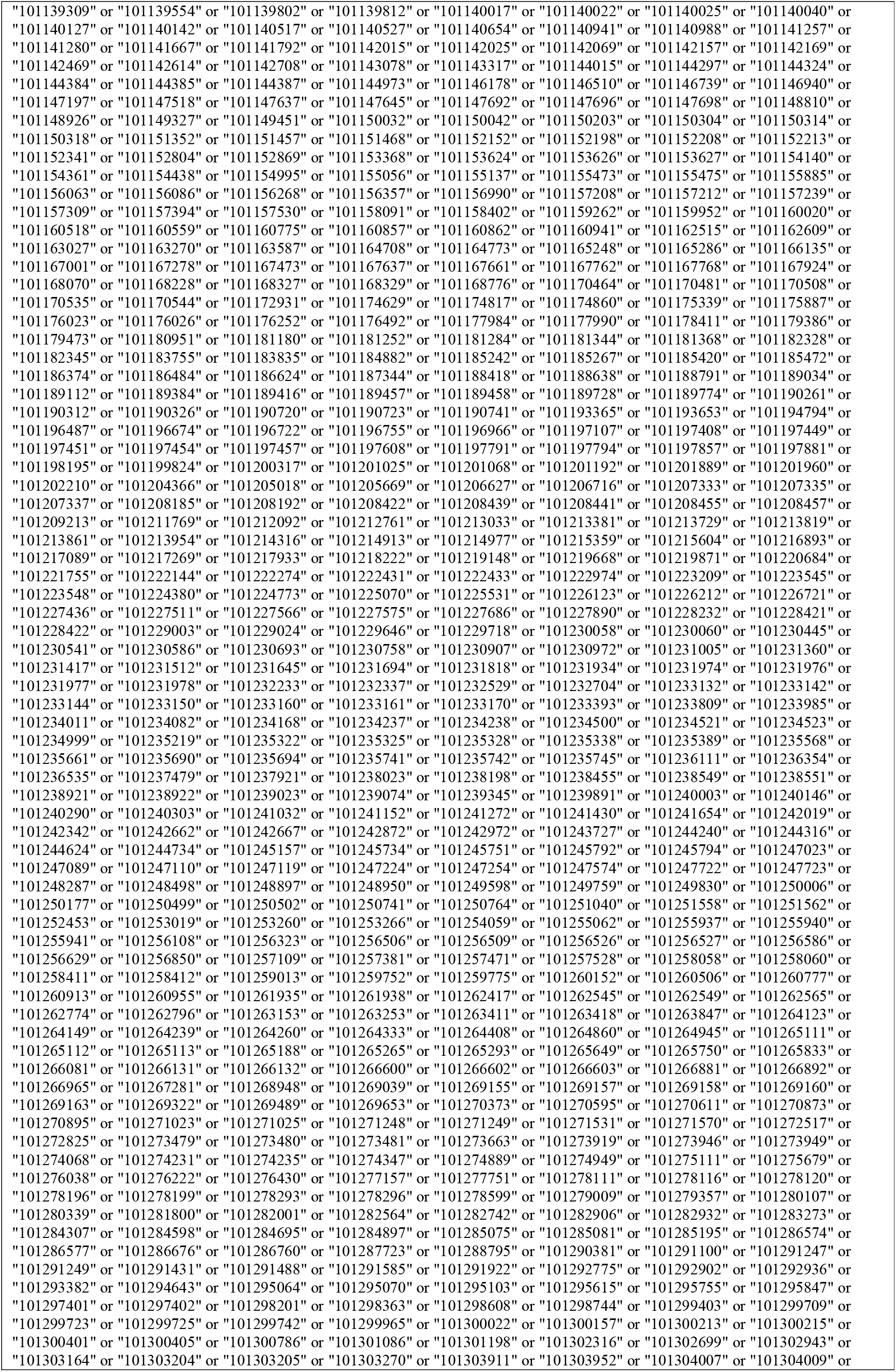
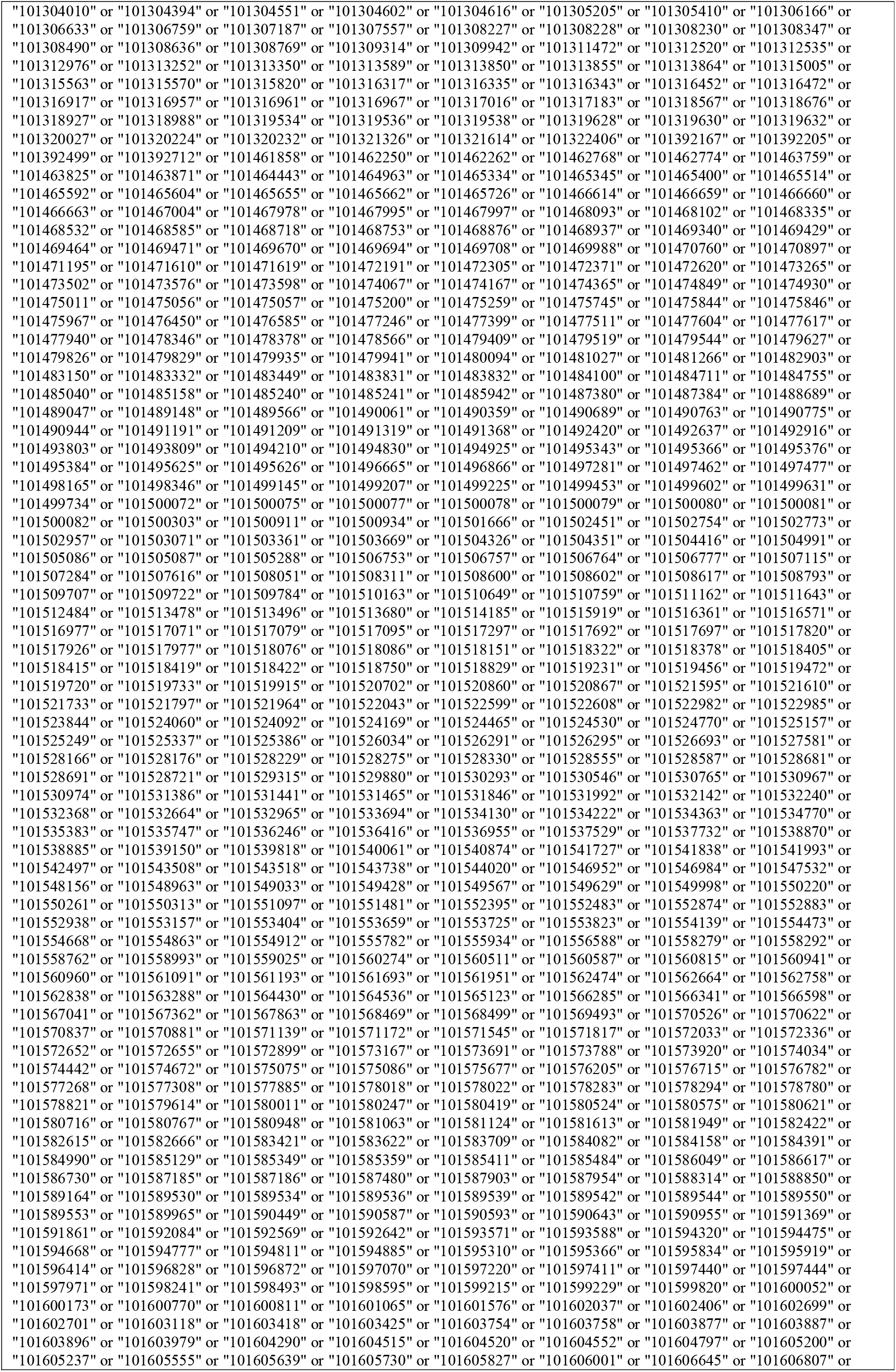
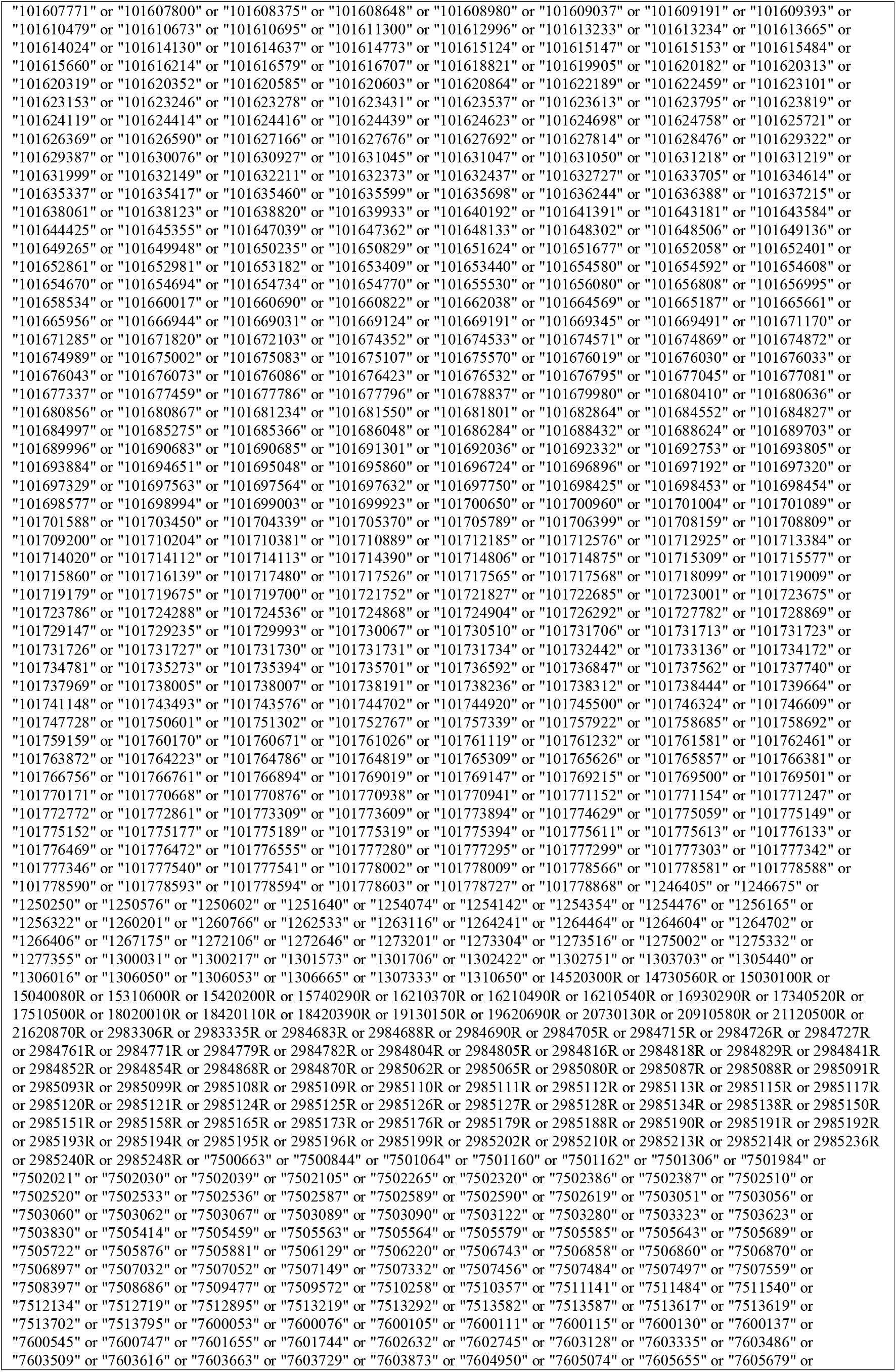
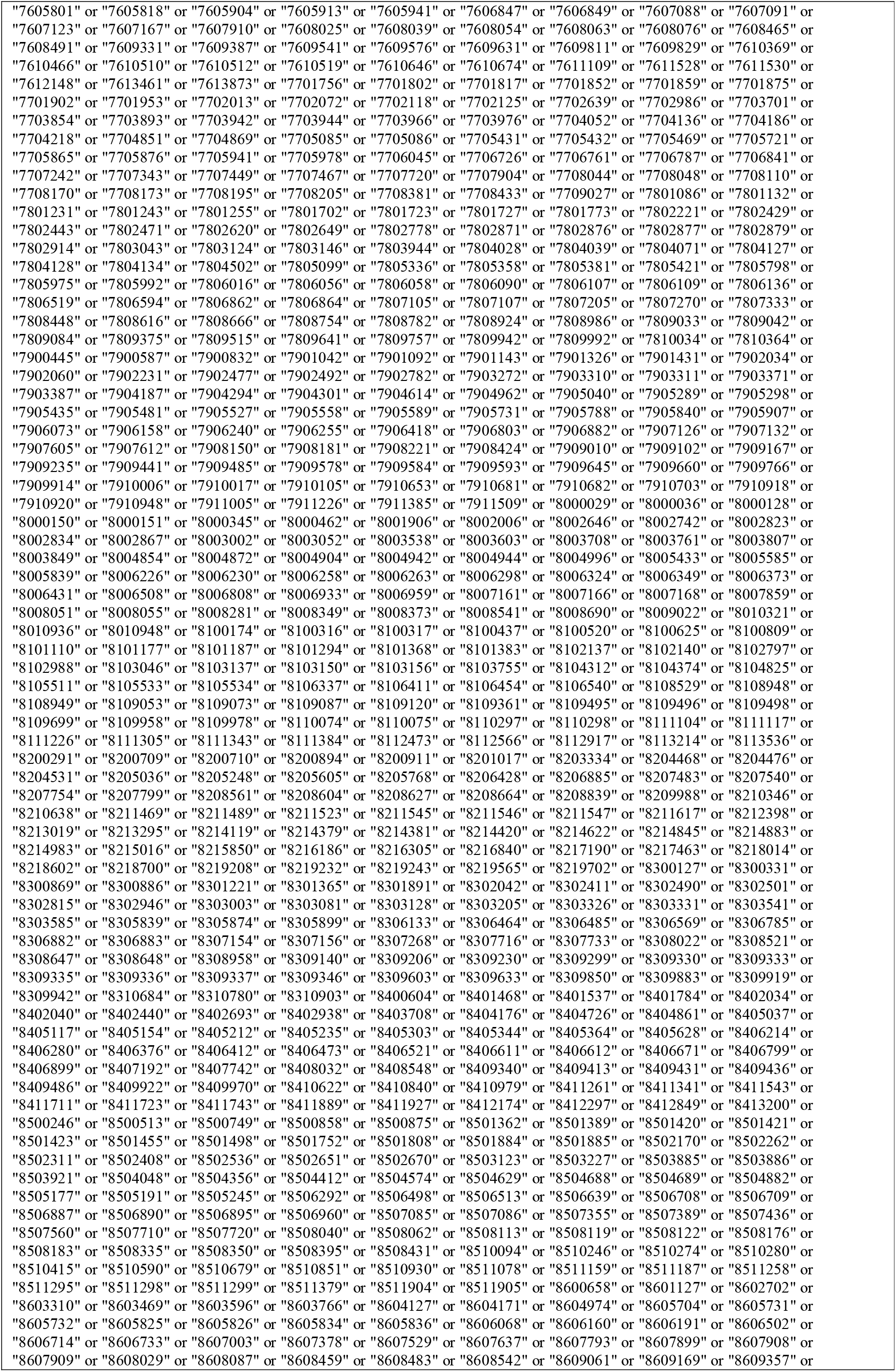
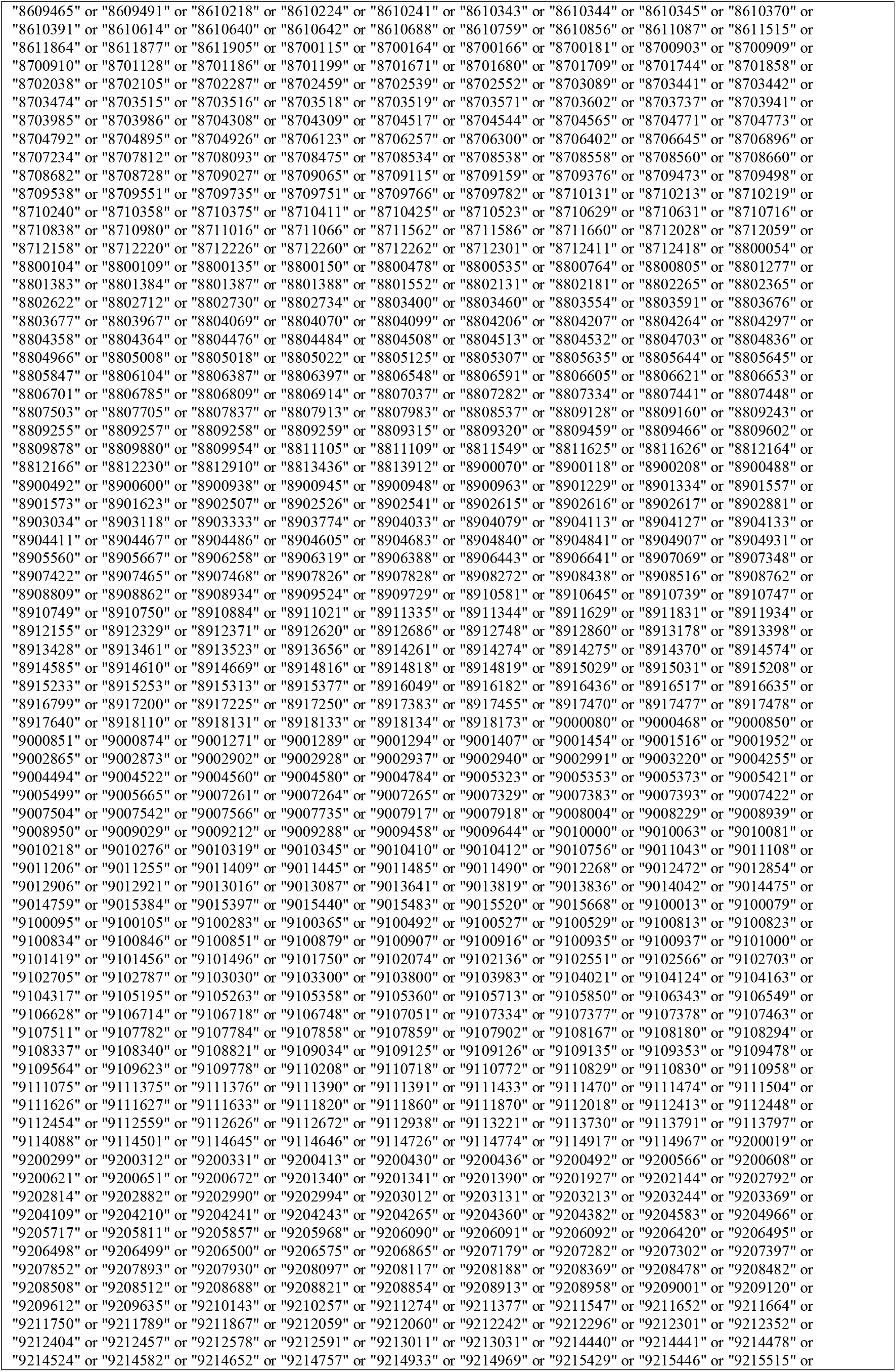
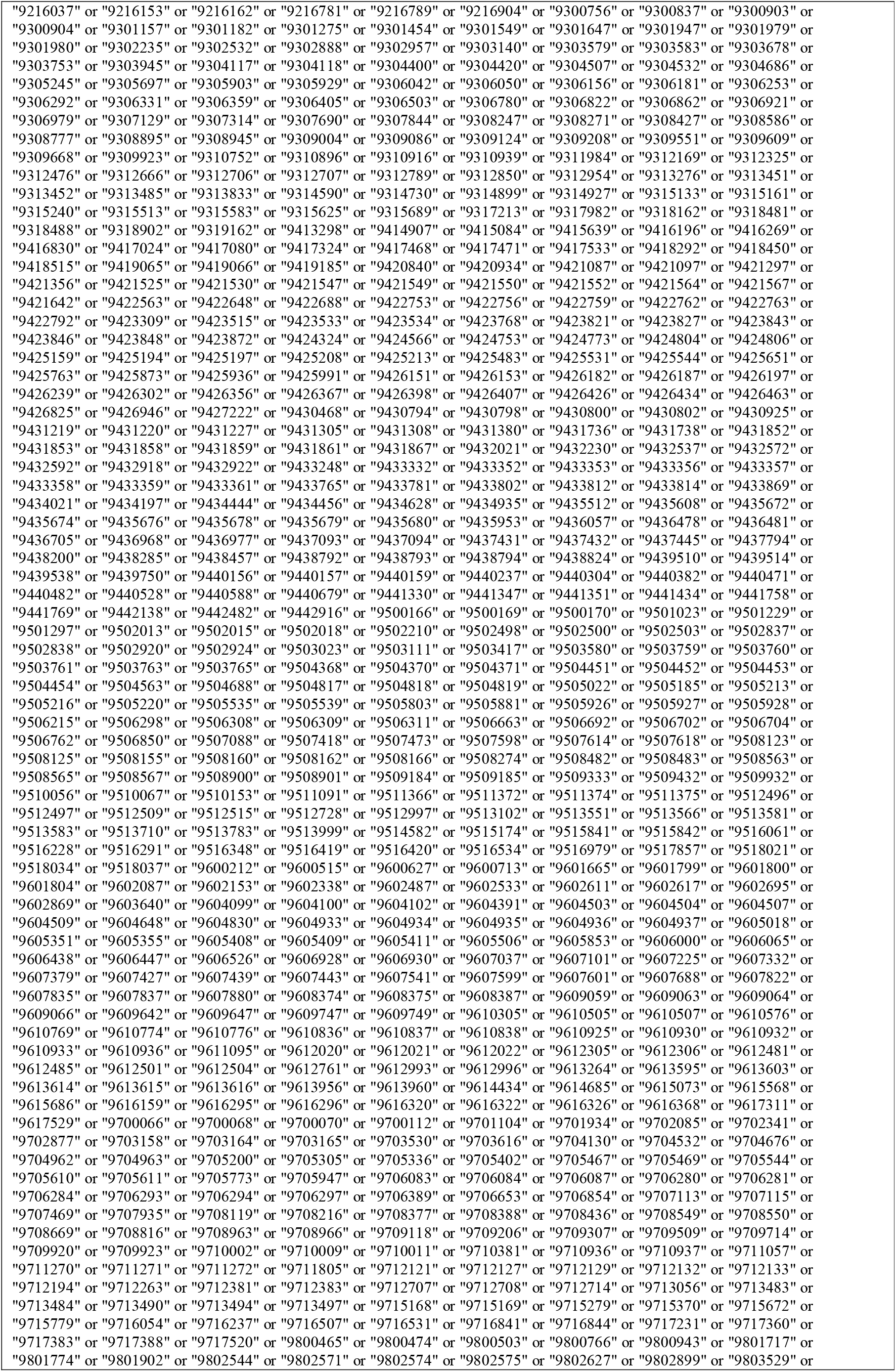
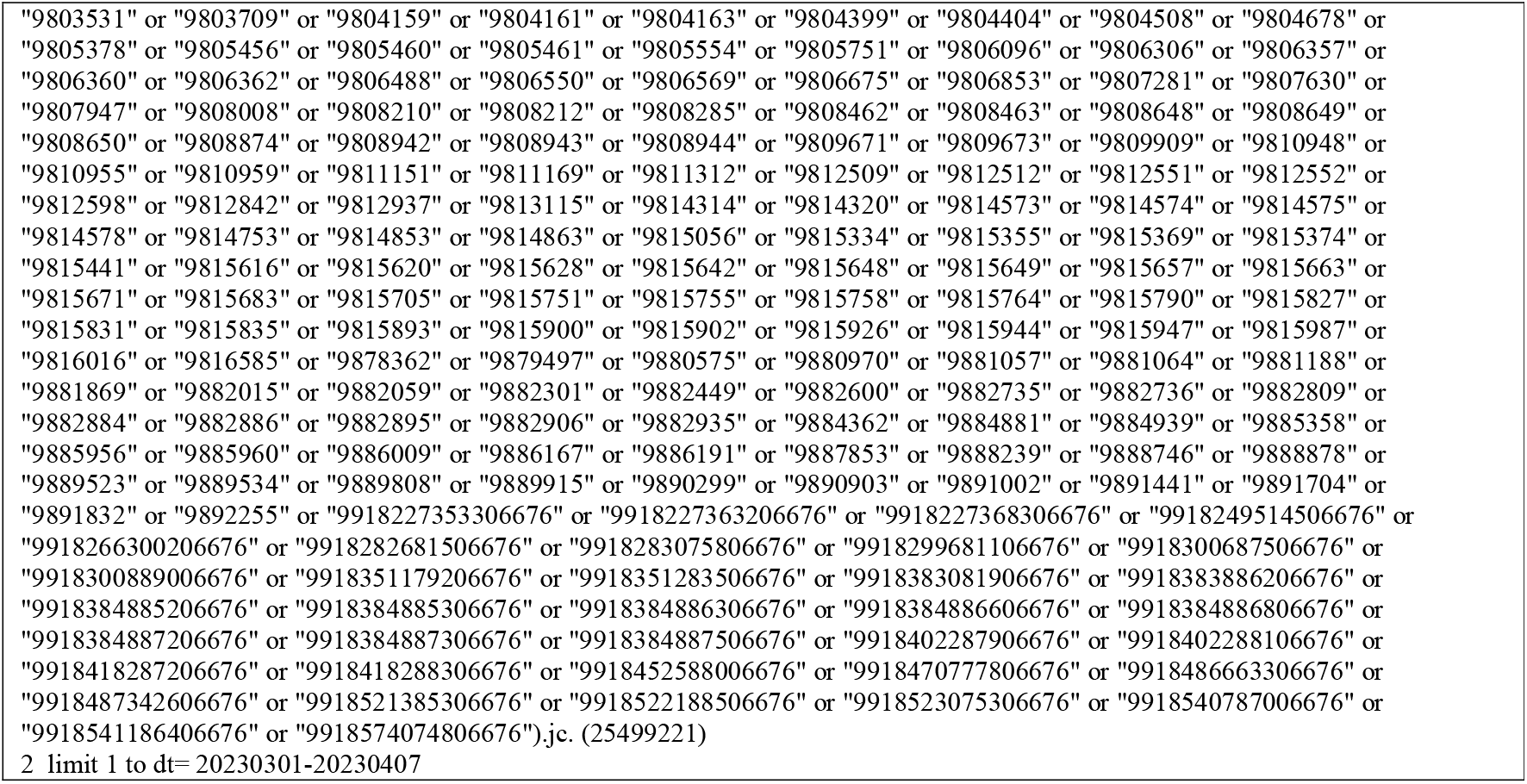

## Appendix 2: Survey Questions

### Initial Screening Question

IMPORTANT NOTE: This survey is intended for participants who are medical researchers (of any kind, whereby your research in one way or another contributes to the field of medicine) who have completed at least one terminal degree in their respective field of study (e.g., PhD or equivalent in your respective field, MD or equivalent in your respective profession) OR have >5 years of experience in a research-focused role (e.g., research coordinator). If you do not meet these eligibility requirements (e.g., student with minimal research experience), you are ineligible to participate.

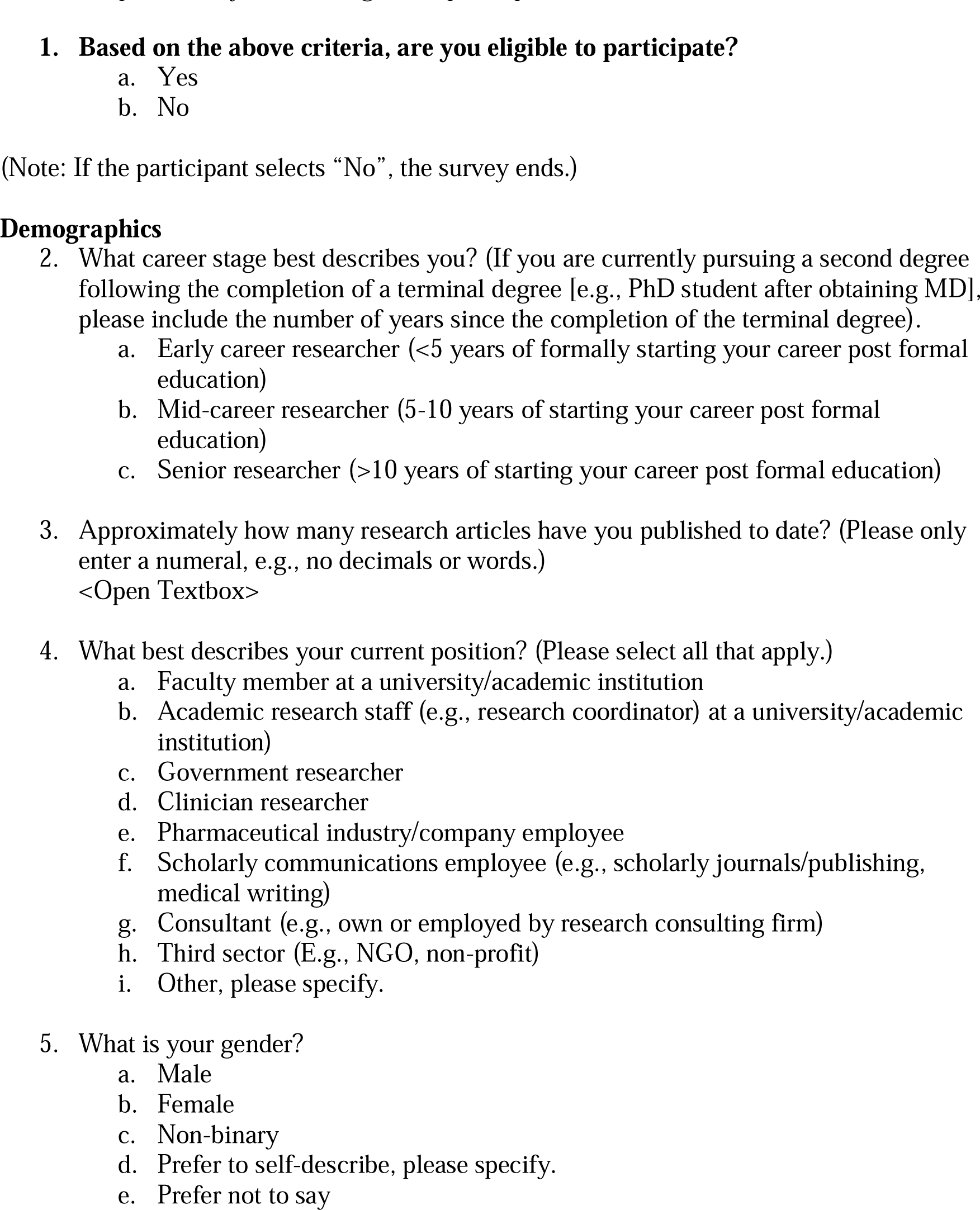
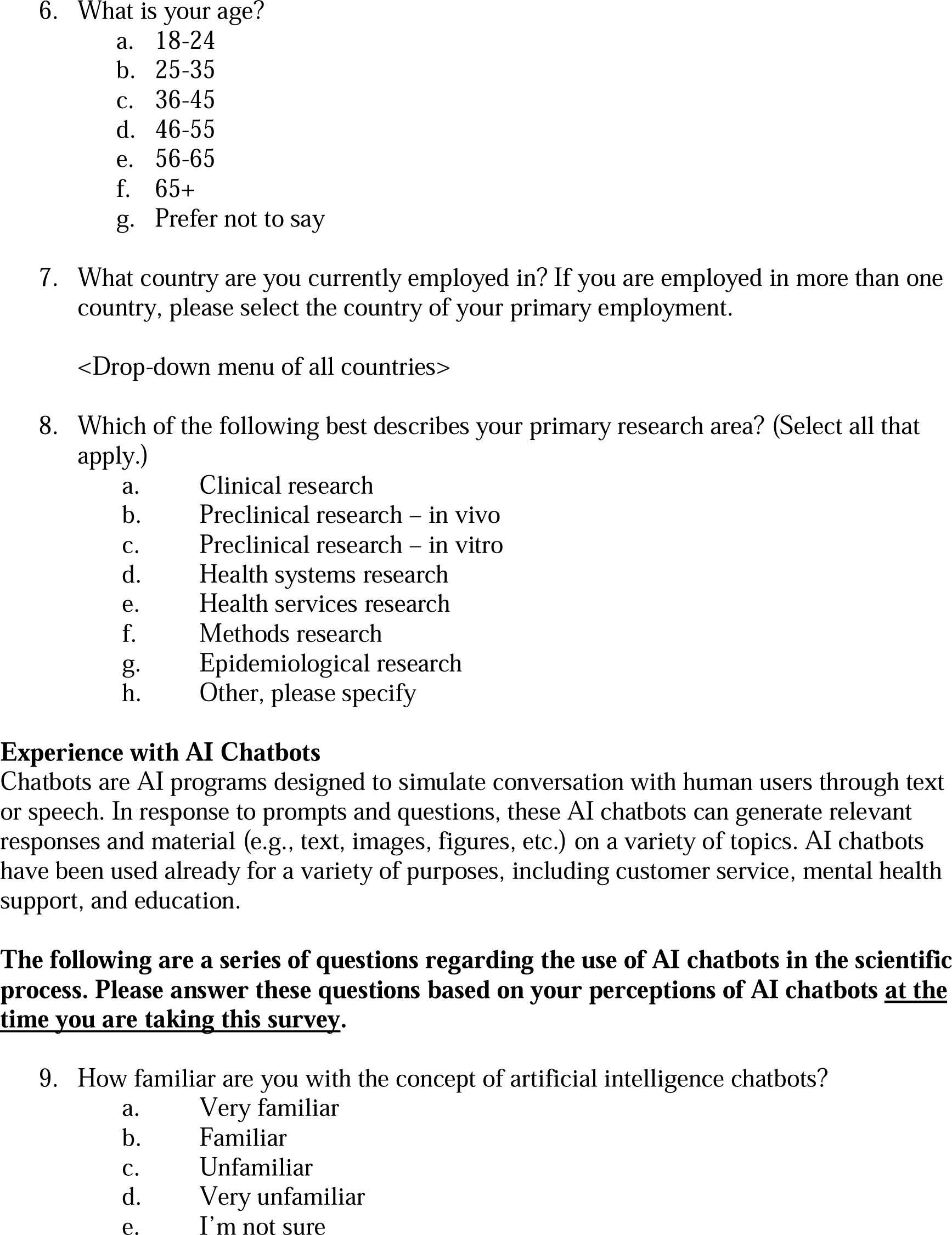
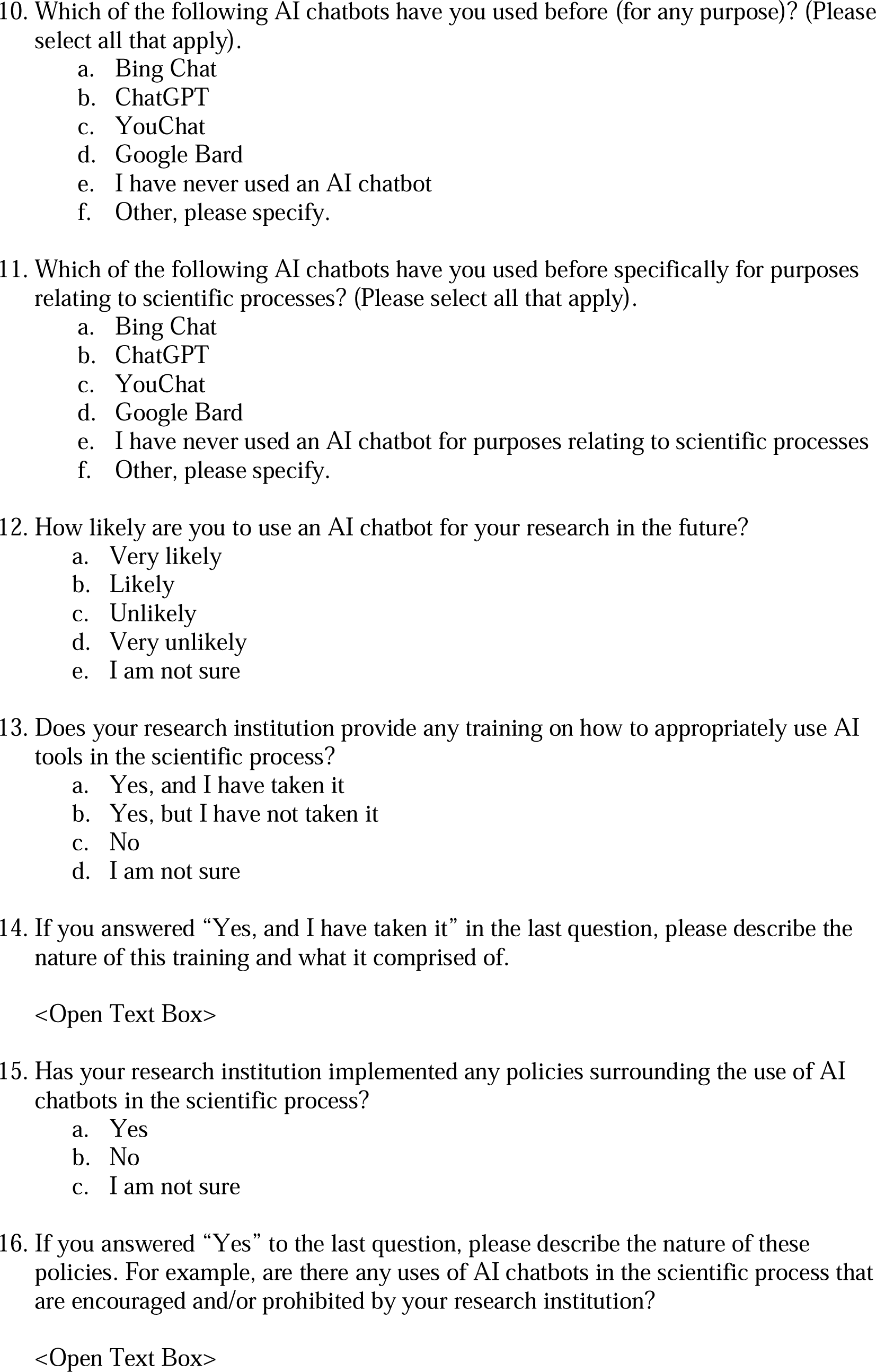
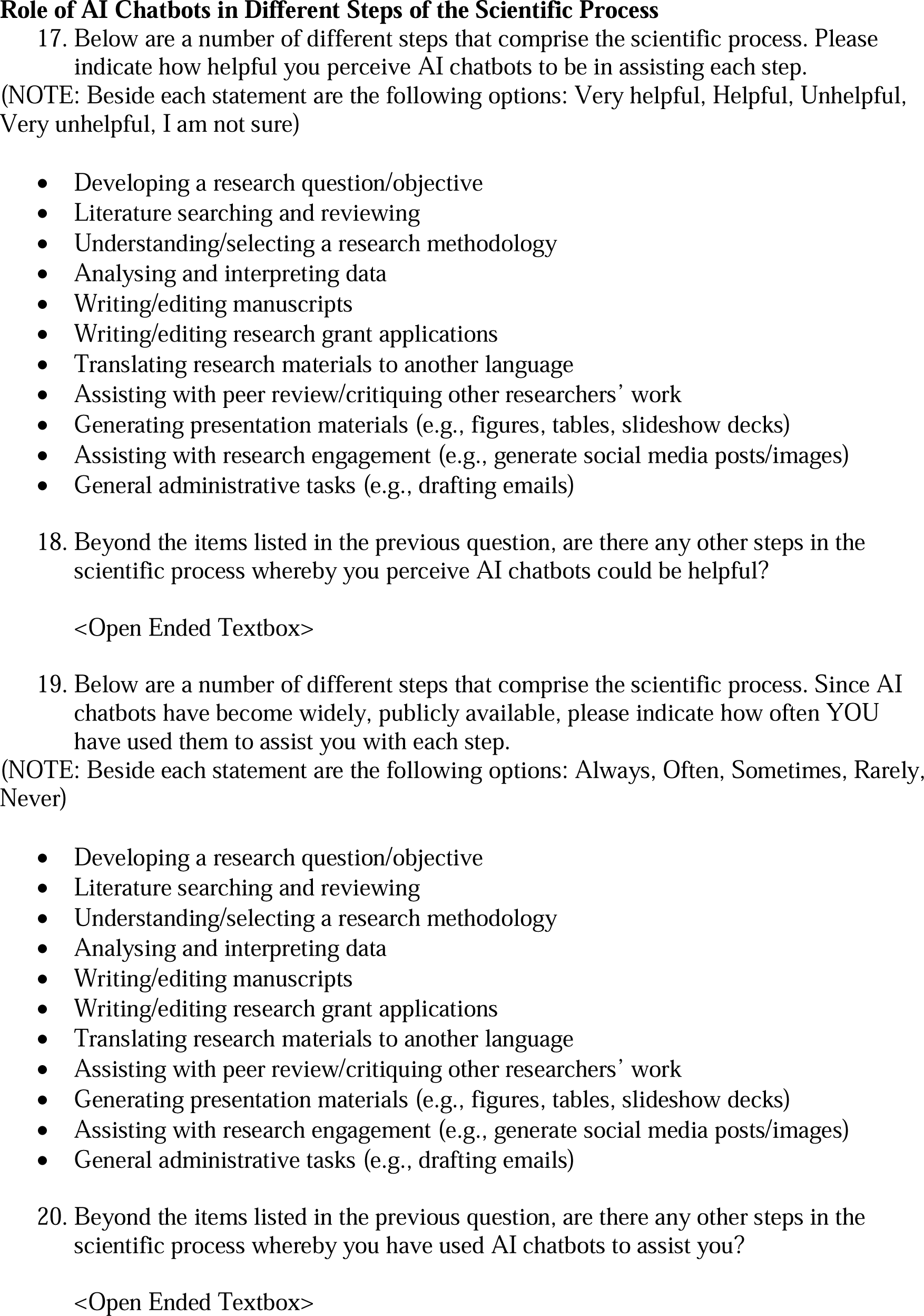
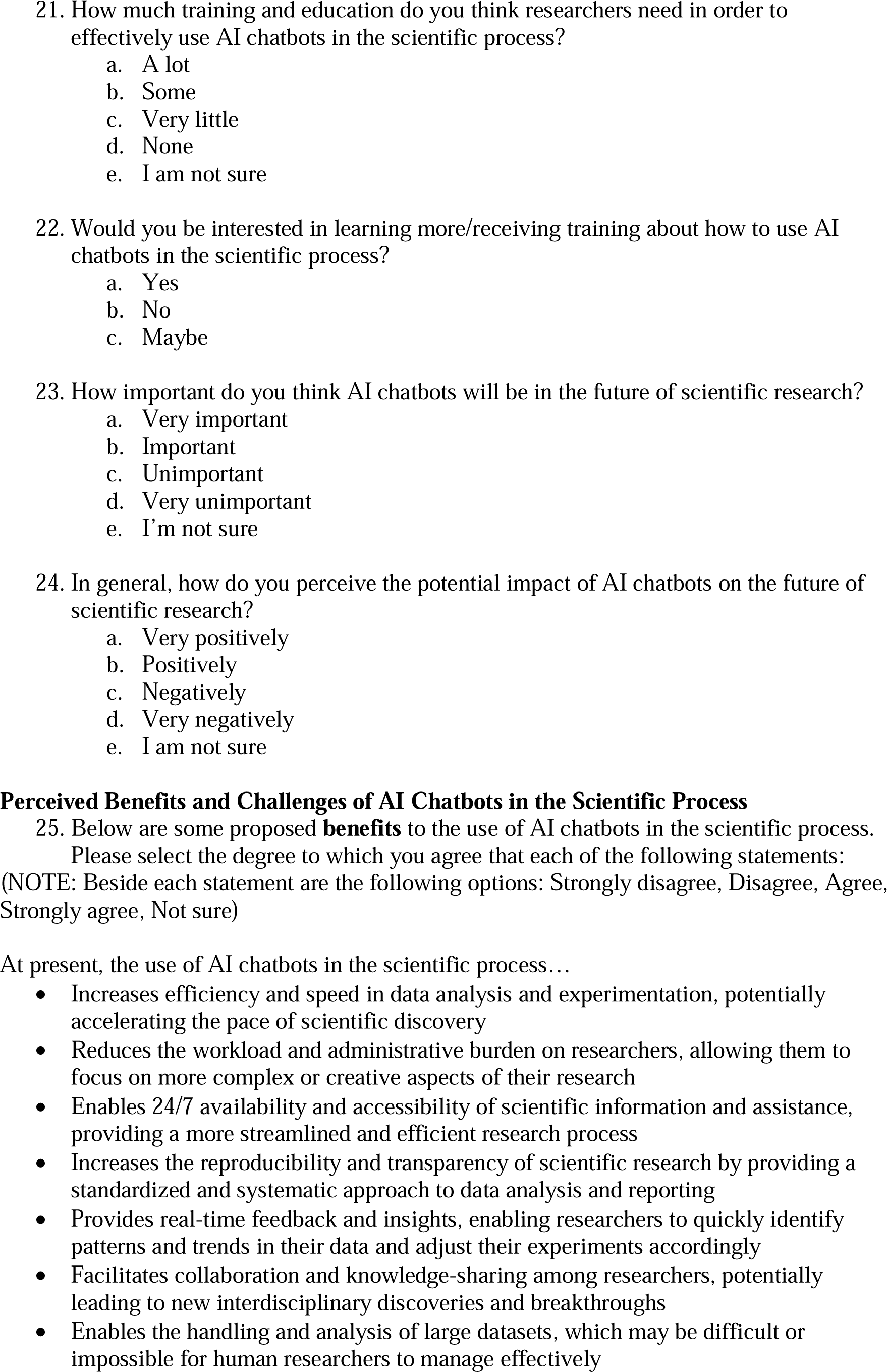
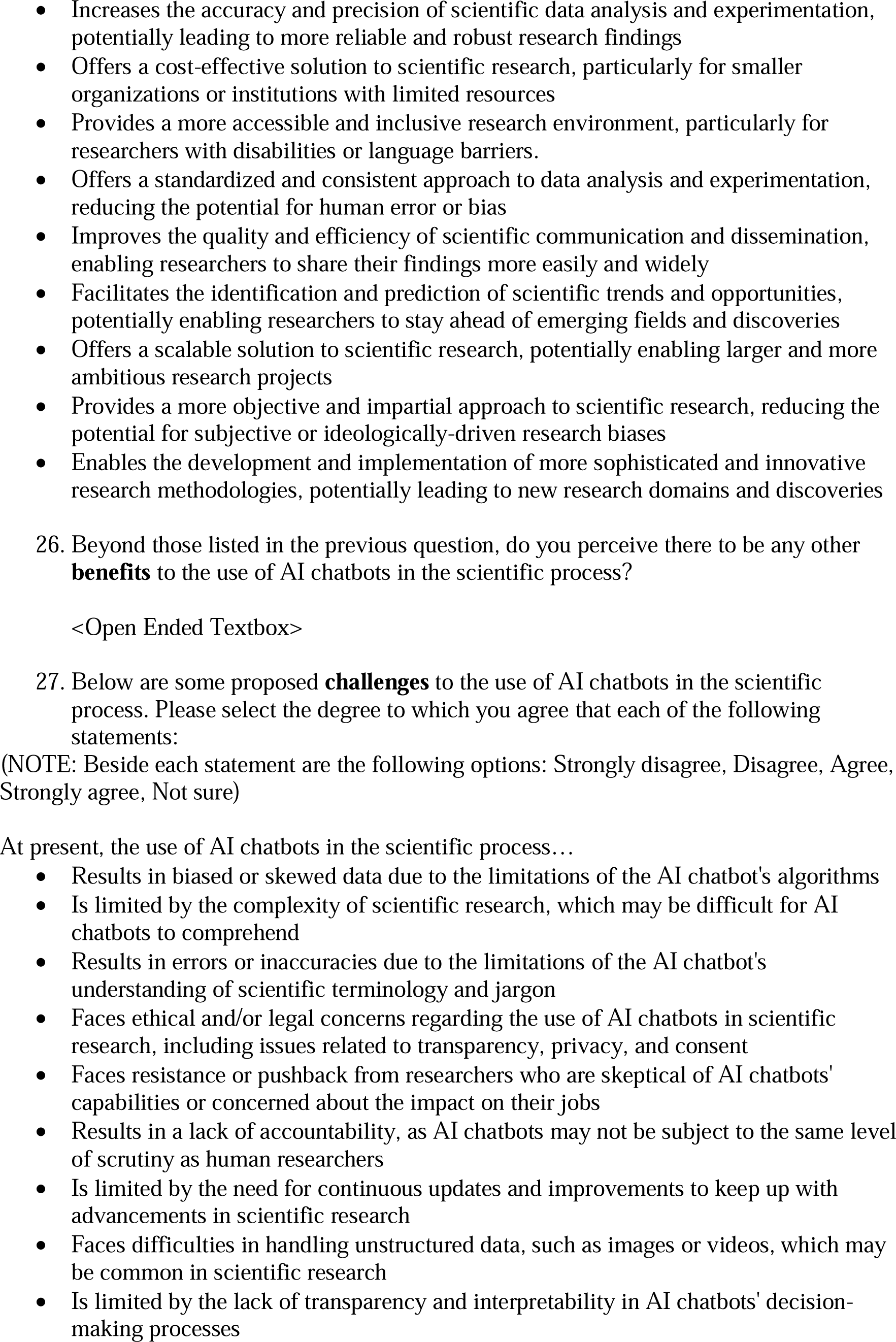
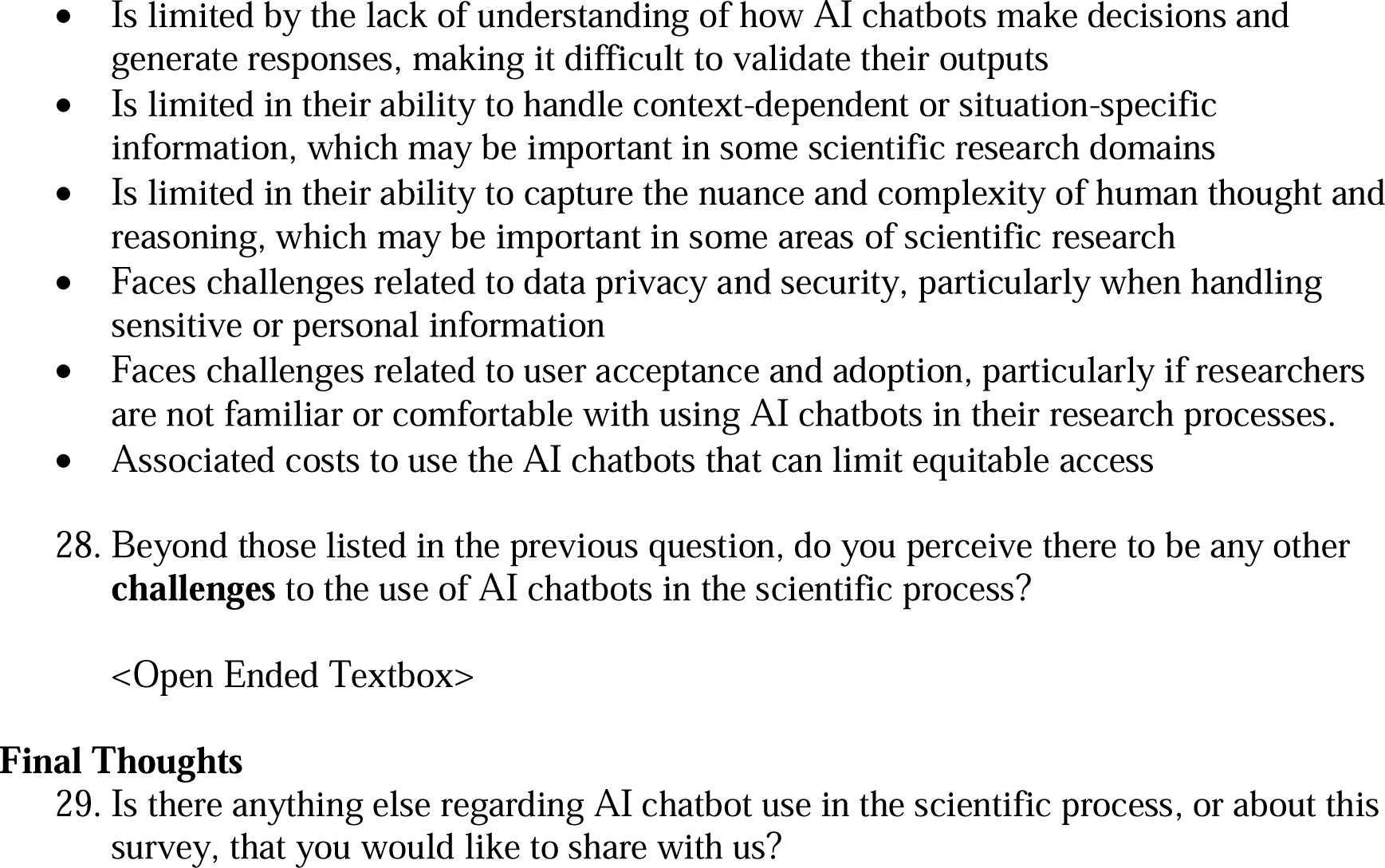

